# Neuroanatomical Features of NAA10- and NAA15-Related Neurodevelopmental Syndromes

**DOI:** 10.1101/2024.06.24.24309433

**Authors:** Rahi Patel, Rikhil Makwana, Carolina Christ, Elaine Marchi, Nathaniel Ung, Randie Harpell, Christina Y. Miyake, Andrea L. Gropman, Gholson J. Lyon, Matthew T. Whitehead

## Abstract

**Background:** *NAA10*-related (Ogden Syndrome) and *NAA15*-related neurodevelopmental syndromes present with varying degrees of intellectual disability, hypotonia, congenital cardiac abnormalities, seizures, and delayed speech and motor development. While there is much data on the clinical manifestations of these conditions, there are few radiologic reports describing the neuroanatomical abnormalities present on imaging.

**Objective:** Our goal was to provide neuroimaging analyses for a subset of probands with *NAA10-* and *NAA15*-related neurodevelopmental symptoms and assess severity, common radiologic anomalies, and changes over time to better understand the pathophysiology of these disease processes.

**Materials and Methods:** Neuroimaging studies from 26 probands (18 with pathogenic variants in *NAA10*, 8 with pathogenic variants in *NAA15*) were collected and analyzed. Size of the cerebrum, brainstem, and cerebellum, as well as myelination, brain malformations, globus pallidus hyperintensity, brain lesions, 4th ventricle size, tegmentovermian angle, cisterna magna size, pituitary size, olfactory tract, palate arch, and choroid plexus abnormalities were analyzed. In depth medical histories were also collected on all probands, including genetic testing results and social, cognitive, and developmental history. The Vineland 3 Adaptive Behavior Scale was also administered to the parents to assess functional status of the probands.

**Results:** On average, individuals with Ogden Syndrome had 5.7 anatomical abnormalities (standard deviation (SD) = 3.0), whereas those with NAA15 related neurodevelopmental syndrome had 2.8 (SD = 2.3) (p = .02). Probands who had more anatomical abnormalities tended to score worse on Vineland assessments, suggesting a possible correlation between the two. Structural-functional anatomic differences seen were preserved such that individuals with greater defects on, for example, motor regions of their scans tested worse on motor portions of the Vineland. Probands followed longitudinally demonstrated several changes between scans, most commonly in the cerebellum, brainstem, and degree of myelination. Such changes were only observed for probands with *NAA10* variants in our cohort.

**Conclusion:** Despite clinical imaging being reported as being predominantly “normal” during routine clinical care, this analysis of a cohort of patients with *NAA10*-related (Ogden Syndrome) and *NAA15*-related neurodevelopmental syndrome by one neuroradiologist has established a range of subtle abnormalities. We hope these findings guide future research and diagnostic studies for this patient population.

## INTRODUCTION

The N-acetyltransferase A (NatA) complex functions as an N-terminus Ala, Val, Ser, Thr, and Cys residue acetyltransferase [1]. It is composed of the N-α-acetyltransferase 10 (NAA10) catalytic subunit and the NAA15 auxiliary subunit and its function is modulated when expressed with several other auxiliary or chaperone proteins [1–4]. Pathogenic variants of *NAA10* and *NAA15* lead to the syndromes NAA10-related neurodevelopmental syndrome, colloquially known as Ogden Syndrome, and NAA15-related neurodevelopmental syndrome, respectively. NAA10-related neurodevelopmental syndrome is primarily X-linked in inheritance, leading to more severe manifestations in males. While there are some similarities between these diseases, such as variable developmental delay [5–10], Ogden syndrome is usually much more severe, possibly due to variants impacting the enzyme domain of NAA10 directly, in comparison to variants in NAA15, where only one copy of NAA15 is affected, as *NAA15* is an autosomal gene. Another possibility is that the NAA10 catalytic subunit might have functions outside of the NatA complex [11, 12]. Both NAA10 and NAA15-related neurodevelopmental syndromes are associated with intellectual disability, autism, dystonia, and congenital heart disease [13–19]. Ogden Syndrome also presents with various cardiac, neurologic, anatomical, and developmental manifestations that are dependent on the specific pathogenic variant of the individual [1, 5–7, 10–12, 20–32].

Intellectual disability is a common neurologic sequela of these variants, with the NAA10 cohort also sometimes developing epilepsy [5, 6]. Despite this, there are few radiologic or histopathologic reports on the neuroanatomical abnormalities present in these syndromes. Ogden Syndrome has been reported to be associated with enlarged ventricles, ocular globe abnormalities, and cerebral dysgenesis [5, 8]. Abnormal MRIs have also been reported in individuals with NAA15-related neurodevelopmental syndrome [13]. This study aims to add to the current body of literature describing the presence or lack of characteristic neuroanatomic variations associated with NAA10 and NAA15-related neurodevelopmental syndromes and correlate their significance to adaptive behavior scores to contribute to better characterization of the disorders and the development of further diagnostic criteria.

## METHODS AND MATERIALS

Neuroimaging studies from 26 probands (18 with pathogenic variants in NAA10, 8 with pathogenic variants in NAA15) were collected from families or from medical facilities, following a secure video conference interview or in-person assessment. Thorough medical histories were collected on all probands, including genetic testing results and social, cognitive, and developmental history. Pathogenicity of the variants was confirmed by clinical exome sequencing. One patient, proband 3 (VI-10), was first described in a 2014 paper by Esmailpour et al^25^. Thus, this proband was not directly interviewed, but medical information and imaging was retrospectively collected and reviewed. Patient 25 was an NAA10 mosaic (76% mutant allele). More information about this patient is available in **Supplementary Note 1.** All supplementary information is available upon request for the MedRxiv preprint.

### Imaging

All imaging was performed during routine clinical care. Usually, participants underwent 1.5 or 3.0 tesla MRI of the brain and spine without the use of intravenous contrast. MR pulse sequences included T1, T2, T2 FLAIR, diffusion weighted imaging and, in some cases, proton density. The neuroimaging consisted of DICOM files obtained from the families or hospitals, which were analyzed in a blinded manner by a fellowship-trained neuroradiologist (M.T.W.) with American Board of Radiology subspecialty certificate in neuroradiology and 12 years of clinical experience after board certification. Imaging studies were qualitatively examined for all visible abnormalities involving the brain and its coverings, head and neck, and spine. We used the term “brain parenchymal volume loss” to encompass both hypoplasia (underdevelopment) and atrophy (wasting). Cerebellar hypoplasia was diagnosed according to definitions of cerebellar atrophy and hypoplasia, as defined by Barkovich^33^. Skull base hypoplasia was diagnosed subjectively based on the appearance of the occipital bone relative to the sizes of posterior fossa CSF spaces and parenchyma. Emphasis was placed upon the cerebrum, brainstem, cerebellum, globus pallidus, pituitary, olfactory bulb, palate, ventral tegmental area, ventricles, and choroid plexus. Additionally, myelination, malformations, and lesions were assessed. Anatomical regions were described based on their size (normal, increased, or decreased) and the severity of the changes (mild, moderate, or severe) based on a combination of objective two-dimensional measurements compared to published aged-matched normative data (fronto-occipital diameter, corpus callosum thickness, brainstem diameter, height and AP diameter of the vermis) and subjective measurements (based on clinical expertise), respectively [33–36].

### Cognitive Assessment

Twenty-one probands (16 NAA10, 5 NAA15) were administered the Vineland Adaptive Behavioral Scales assessment, third edition (Vineland). The Vineland is composed of three core domains—Communication, Daily Living Skills, and Socialization—with an optional fourth motor domain. These core domains are each divided into three sub-domains, while motor is divided into two sub-domains. A composite score (ABC) is then calculated based on the overall results. The Vineland was administered by two trained assessors at various points in the probands’ lives. Vineland is not officially validated for use in individuals under the age of eight. However, to better characterize the motor components of the disease, it was administered to all participants regardless of their age, as long as they were able to perform the minimum tasks required for assessment. All probands with any Vineland scores were included in the analysis.

### Analysis

Anatomical features and severity were coded numerically and tabulated in an Excel spreadsheet and combined with the Vineland data. Single tailed ANOVA were calculated comparing specific anatomic abnormalities with correlated Vineland domain data. Variations in cortical volume loss and severity of cortical malformations were compared against the composite ABC score. Cerebellar and globus pallidus abnormalities were compared against the motor domain. Variation in white matter were compared against all Vineland domains due to its importance across all brain functions. Variations in cortical volume loss were then clustered together such that those with normal or mild cortical thinning were grouped and those with moderate or marked thinning were grouped. These were then compared against participants who were classified as mildly dysfunctional (corresponding to Vineland scores of 100-50) and those who were classified as severely dysfunctional (scores of 49-0). While the Vineland manual provides ranges to help designate categories for levels of adaptive behavior, the study sample was too low functioning to match their categories. Instead, clusters were formed to help increase power due to low small sample sizes. Fisher exact tests were then performed to compare associations of cortical thinning, cerebellar volume, and globus pallidus hyperintensities and categories of clinical dysfunction as defined by Vineland scores. Two tailed, unequal variance t-tests were also performed between individuals with NAA10-related neurodevelopmental syndrome and NAA15-related neurodevelopmental syndrome to assess differences between groups. Alpha was set at .05 for all calculations except for the white matter comparisons with the Vineland functional domains. This ANOVA included 16 comparisons (one per Vineland domain and subdomain), necessitating a Bonferroni correction to reduce the risk of false positives arising from multiple sequential comparisons using the same data. The modified alpha for the white matter calculation was set at .003 (m = 16).

Participants who were imaged longitudinally were also compared to their previous scans. Neuroanatomical deficits were then correlated with the probands’ behavior based on in-depth interviews conducted by GJL that corresponded with the approximate time of imaging.

## RESULTS

Neuroimaging from a total of 26 probands with either Ogden Syndrome (n=18) or NAA15-related neurodevelopmental syndrome (n=8) ranging in age from 0 to 40 years old was analyzed, with several probands being analyzed longitudinally. A demographic breakdown of the probands included can be seen in **Table 1**. Full mutation information by proband number can be viewed in **Supplemental Table 1.**

**Table 1.** Demographic Information for NAA10 and NAA15 Probands.

| Characteristics | NAA10 (n=18) | NAA15 (n=8) |
| --- | --- | --- |
| **Sex** |  |  |
| Male | 5 | 2 |
| Female | 13 | 6 |
| **Race** |  |  |
| Caucasian | 16 | 8 |
| Asian | 1 | 0 |
| African American | 1 | 0 |
| **Ethnicity** |  |  |
| Hispanic | 1 | 0 |
| Non-Hispanic | 17 | 8 |
| **Age Range** | 0 years - 40 years | 1 year - 15 years |
| **Country of Residence** |  |  |
| USA | 12 | 6 |
| Australia | 1 | 2 |
| United Kingdom | 2 | 0 |
| Finland | 1 | 0 |
| Austria | 1 | 0 |
| China | 1 | 0 |

On average, probands with Ogden Syndrome had 5.7 anatomical abnormalities (standard deviation (SD) = 3.0) and those with NAA15 related neurodevelopmental syndrome had 2.8 (SD = 2.3). This difference was significant (p = .02). Individuals with Ogden Syndrome were also significantly more likely to experience decreased cortical volume (p = .01), hyperintensities of the globus pallidus (p = .02), and abnormalities of the olfactory bulb (p = .004), as compared to those with NAA15 related neurodevelopmental syndrome. A breakdown of the specific neuroradiologic abnormalities observed in the NAA10 and NAA15 cases can be seen in **Table 2**. Full patient neuroimaging data is available in **Supplemental Table 2**.

**Table 2.** Summary of Neuroanatomical Variations Observed in NAA10 versus NAA15 Related Neurodevelopmental Syndromes in Most Recent Scan. *Asterisk indicates that imaging finding switched from another result on a previous scan.

| ***Brain Structures affected*** | **NAA10 (n=18)** | **NAA15 (n = 8)** |
| --- | --- | --- |
| ***Cerebrum volume*** |  |  |
| Normal | 3 | 6 |
| Cerebral white matter hypoplasia and/or diffuse volume loss | 12 | 2 |
| Cerebral white matter hypoplasia and/or posterior-predominant volume loss | 2 | 0 |
| Cerebral volume loss +/- hypoplasia | 1 | 0 |
| ***Brainstem volume*** |  |  |
| Normal | 9 | 5 |
| Diffuse volume loss and/or hypoplasia | 3* | 0 |
| Midbrain volume loss and/or hypoplasia | 1 | 0 |
| Midbrain and pons volume loss and/or hypoplasia | 1 | 0 |
| Pons volume loss and/or hypoplasia | 4* | 3 |
| Medulla volume loss and/or hypoplasia | 0 | 0 |
| ***Cerebellum Volume*** |  |  |
| Normal | 8 | 7 |
| Diffuse volume loss and/or hypoplasia | 4* | 0 |
| Volume loss | 1 | 0 |
| Hypoplasia | 0 | 0 |
| Vermis hypoplasia | 2* | 0 |
| Vermis volume loss | 0 | 1 |
| Hemisphere hypoplasia | 3* | 0 |
| Hemisphere volume loss | 0 | 0 |
| ***Myelination*** |  |  |
| Normal | 15* | 8 |
| Delayed and/or hypomyelination | 3 | 0 |
| ***Malformation*** |  |  |
| None | 17 | 6 |
| Mild gyral simplification | 1 | 1 |
| Malformations of cortical development (MCD) | 0 | 1 |
| ***Globus Pallidus Hyperintensity*** |  |  |
| No | 13* | 8 |
| Yes | 5 | 0 |
| ***Brain Lesions*** |  |  |
| No | 16 | 6 |
| Yes | 2 | 2 |
| ***Fourth Ventricle Size*** |  |  |
| Normal | 5 | 5 |
| Mild enlargement | 12 | 3 |
| Moderate enlargement | 1 | 0 |
| ***Tegmento-Vermian Angle (TVA)*** |  |  |
| Normal | 8 | 5 |
| Mild increase | 10 | 3 |
| ***Cisterna Magna Size*** |  |  |
| Normal | 4 | 5 |
| Mild enlargement | 14 | 3 |
| ***Pituitary*** |  |  |
| Normal | 16 | 7 |
| Hypoplastic | 2 | 1 |
| ***Olfactory*** |  |  |
| Normal | 11 | 8 |
| Hypoplastic | 7 | 0 |
| ***Palate*** |  |  |
| Normal | 16 | 8 |
| High Arch | 2 | 0 |
| ***Choroid Plexus in Left Ventricle*** |  |  |
| Normal | 13 | 7 |
| Mild Cystic | 5 | 1 |

Abnormalities in brain volume were observed across a spectrum of severity in probands diagnosed with Ogden syndrome, as illustrated in **Figure 1**. Some probands, including patient 13, also had abnormal parenchymal growth over time. This patient had normal cerebellar volume at age [0–4] years, but later scans showed mild hypoplasia of the cerebellar vermis **(Figure 2).**

**Figure 1a-d.**
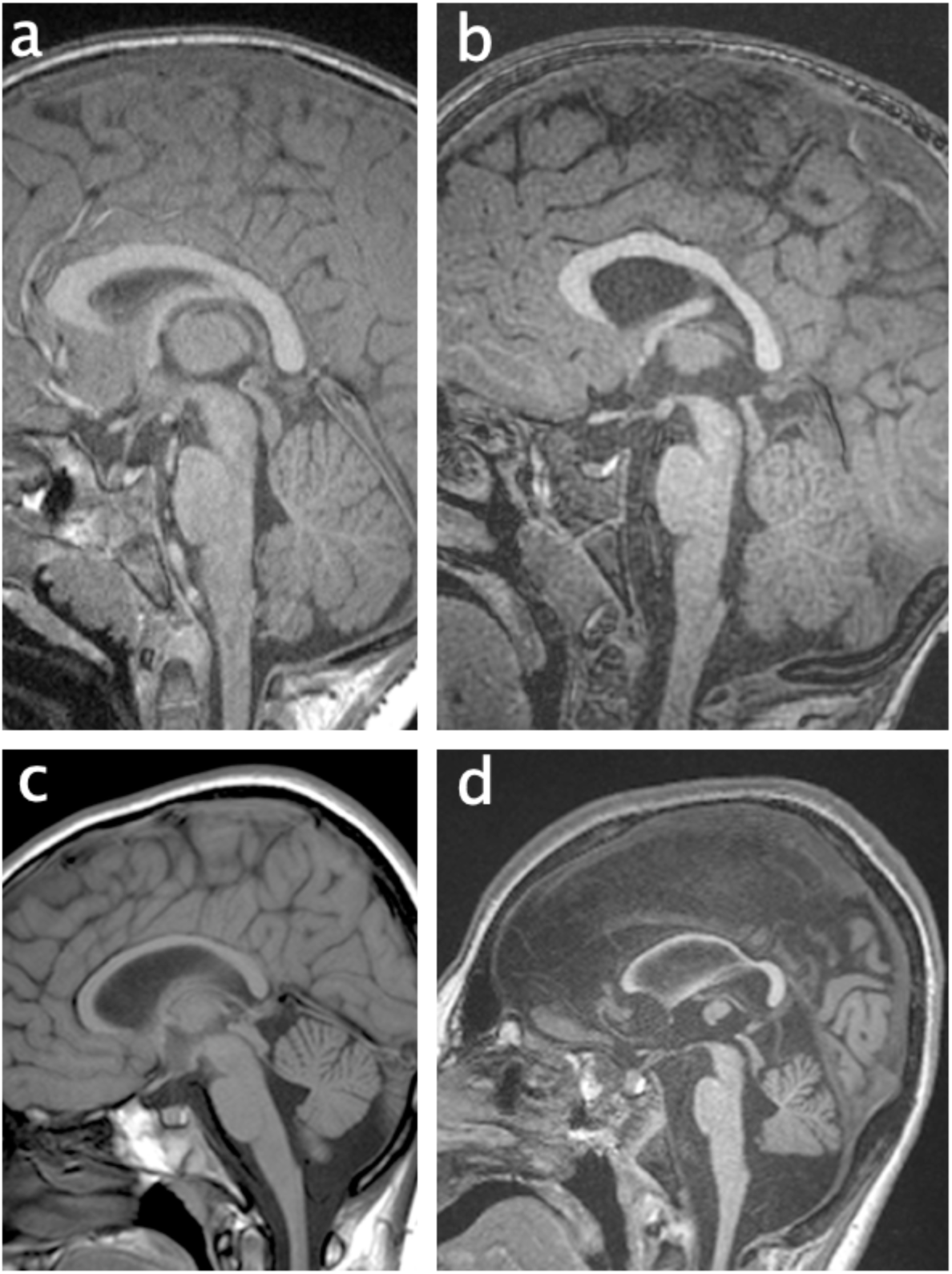
Sagittal midline TI-weighted brain MRI collage demonstrating the phenotypic range of brain volume in 4 different probands with Ogden syndrome. a=normal (patient 15), b=mildly decreased cerebral volume with mild callosal thinning (patient 12), c=mildly diffusely decreased brain volume (patient 2), d=marked cerebral, moderate brainstem, and mild cerebellar volume reduction (patient 9).

**Figure 2.**
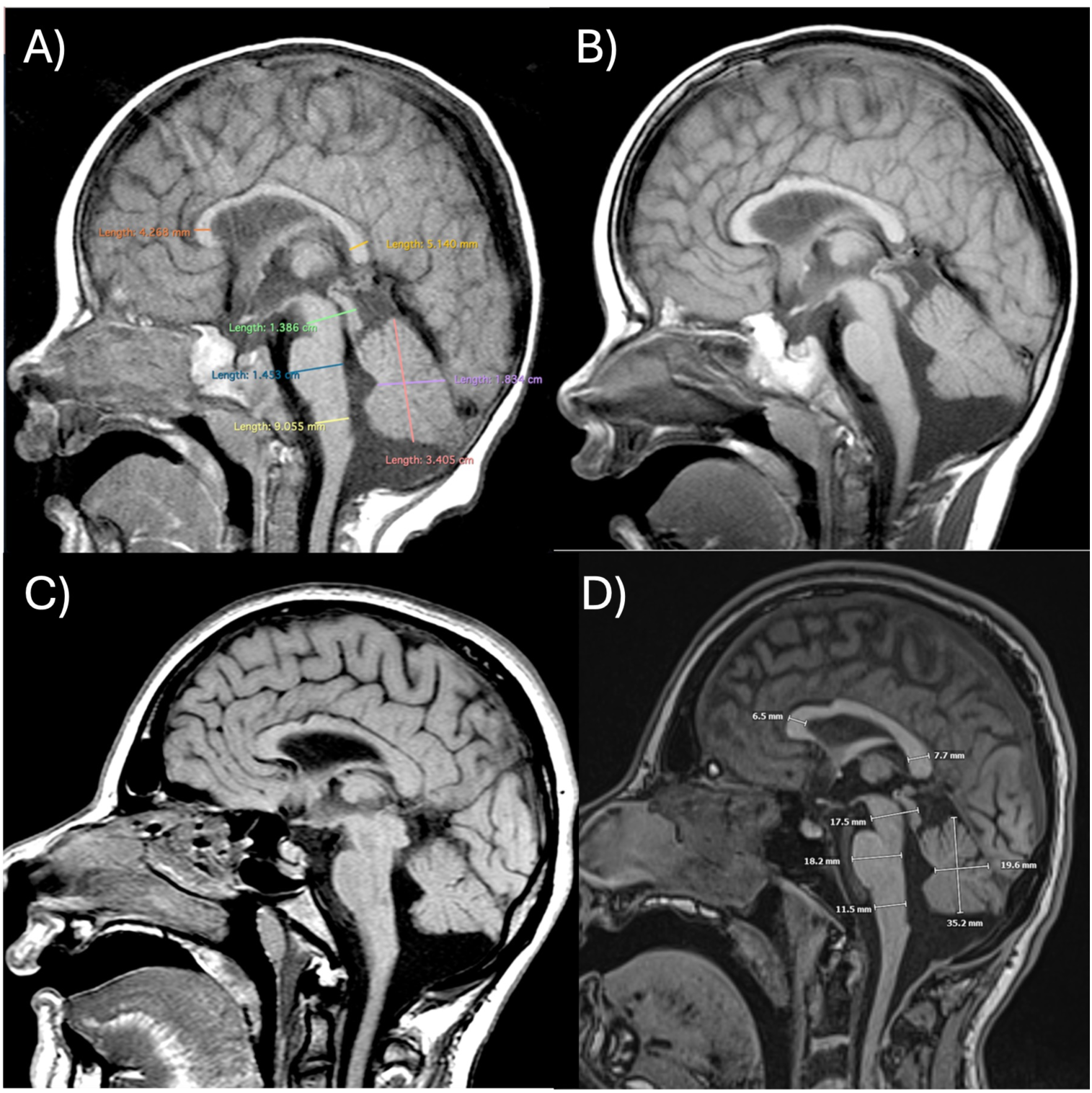
Sequential sagittal midline T1-weighted brain MR images from patient 13 at ages (a) [0–4] years, (b) [0–4] years, (c) [5–9] years, and (d) [15–19] years showing mild cerebral, cerebellar, and brainstem hypoplasia with lack of appropriate parenchymal growth over time.

Globus pallidus signal hyperintensity on T2-weighted MRI was observed in five probands **(Figure 3).** While some globus pallidus signal can be an age-related physiological finding, the degree of hyperintensity in these probands was abnormal for their age.

**Figure 3.**
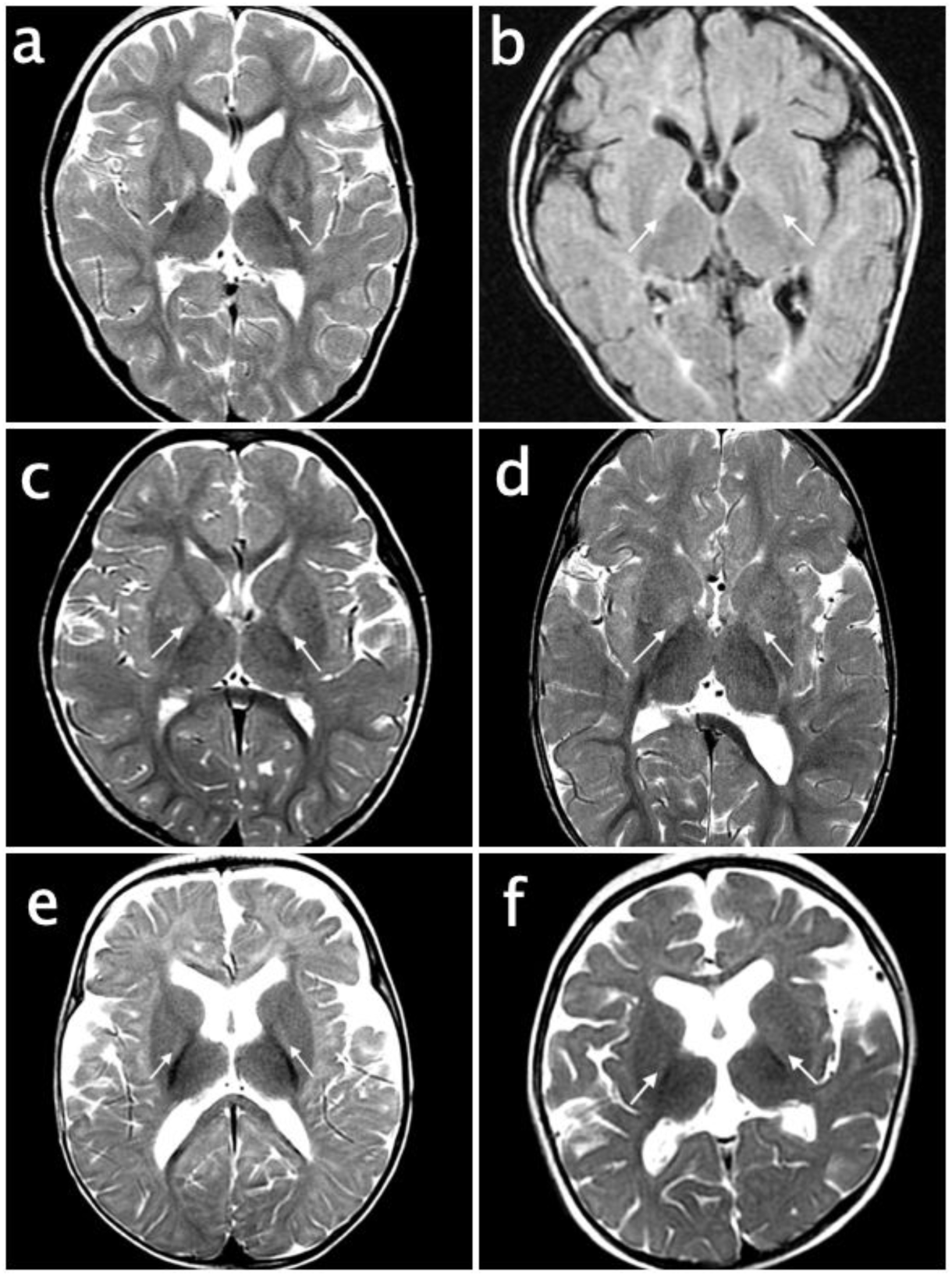
Brain MRI collage showing globus pallidus signal hyperintensity (arrows) on T2WI (a, c-f) and T2/FLAIR (b) in 5 patients with Ogden syndrome. Figures a and b are from the same patient (patient 13) at [0–4] years of age; the T2/FLAIR sequence reveals more widespread myelin deficiency abnormal white matter signal for age (b). Figure c=patient 4 ([0–4] years), d=patient 8 ([0–4] years), e=patient 5 ([0–4] years), figure f=patient 7 ([0–4] years).

Out of the 26 probands, 4 presented with brain lesions visible on imaging. Patient 3 (NAA10) had left posterior frontal corona radiata gliosis at [10–14] years. Patient 8 (NAA10) had mild right posterior frontal deep and peri-atrial parietotemporal gliosis on multiple MRIs. Probands 10 (NAA15) and 11 (NAA15) had multiple punctate hyperintense frontal and parietal white matter lesions and gliosis. There was also variation in the relative size of the 4th ventricle and cisterna magna between probands. These enlargements were observed without concomitant supratentorial ventriculomegaly or dilation of the cerebral aqueduct.

Several probands exhibited changes between scans, as exhibited in **Supplemental Table 3**. Seven NAA10 probands (39%) and 3 NAA15 probands (38%) had more than one scan available for analysis for changes over time. Please refer to **Supplementary Note 2** for more details on individual cases.

Of those with neuroimaging analyzed, 21 (16 NAA10, 5 NAA15) probands had Vineland assessments performed. Individuals with normal cortical volume (n = 5), mild cortical thinning (n = 13), moderate cortical thinning (n = 2), and severe cortical thinning (n = 1) were compared. Those with normal cortical volume had the highest mean ABC score of 60.2 (standard deviation (SD) = 16.2). Those with mild, moderate, and severe decreases in cortical volume had mean scores of 50.6 (SD =21.0), 32.5 (SD = 7.5), and 31, respectively, which correlates with increasing severity. This downward trend was not statistically significant. Individuals with mild adaptive behavioral dysfunction (Vineland ABC scores 100 – 50, n = 11) were compared to those with severe dysfunction (scores 49-0, n = 9). Of those with mild dysfunction, all of them had normal or mild levels of cortical thinning. Of those with severe dysfunction, 6 had normal or mild levels of cortical thinning and 3 had moderate or marked levels of cortical thinning. These differences were not significant (p = .07). Full Vineland results and developmental notes are available in **Supplemental Table 4.**

Probands with normal cerebellar volume (n=8) and mild decreases in cerebellar volume (n=3) were also assessed to identify potential differences in motor Vineland scores. The mean motor domain score for probands with normal cerebellar volume was 51.8 (SD = 22.0). With mild decreases in cerebellar volume, the mean score was 29.7 (SD = 13.7). However, once again, this was not a statistically significant difference. After clustering individuals by adaptive behavioral dysfunction, there were no significant differences between groups with normal or mildly decreased cerebellar volume (p = .2). Those with mild motor dysfunction all had normal cerebellar volume (n = 5). Those with severe motor dysfunction had either normal (n = 3) or mildly decreased (n = 3) cerebellar volume. When comparing motor scores of individuals with absent globus pallidus hyperintensities (n = 4) to those with globus pallidus hyperintensities (n = 7), there was also no significant difference between the scores [mean = 38 (SD = 13.4) versus 33.3 (SD = 16.8), respectively]. After clustering individuals by adaptive behavioral dysfunction, there were no significant differences between groups and the presence of globus pallidus hyperintensities (p = .06). Of those with mild motor dysfunction, 2 had globus pallidus hyperintensities and 5 did not. Of those with severe motor dysfunction, 6 had globus pallidus hyperintensities and 5 did not.

In comparing written sub-domain scores between those with normal white matter (n = 18, mean = 4.8, SD = 3.8) and those without (n = 3, mean = 1, SD = 0), there was a significant difference between the written sub-domain scores (p = .002). No other comparisons showed statistically significant differences.

## DISCUSSION

Both NAA10 and NAA15-related neurodevelopmental syndromes are systemic disorders affecting multiple systems. Shared features include variable neurodevelopmental disability, motor dysfunction, and cardiovascular abnormalities. In addition to being more severe than NAA15-related neurodevelopmental syndrome, Ogden Syndrome is also more strongly associated with craniofacial, gastrointestinal, and metabolic dysfunction, whereas such abnormalities are often absent or less severe in NAA15-related neurodevelopmental syndrome [5, 37].

Further evidence for the increased severity of Ogden Syndrome compared to NAA15 related neurodevelopmental syndrome was showcased by the significantly greater number of neuroanatomical malformations associated with the former than the latter. Cortical thinning has been associated with decreased adaptive functioning in autism spectrum disorder, which could be an explanation for why those with NAA10 related neurodevelopmental syndrome tend to receive lower ABC standard scores, on average, than those with NAA15 related neurodevelopmental syndrome [9, 38, 39]. The findings of lower ABC scores in groups that were considered to have more severe levels of cortical volume loss supports this observation. However, cortical volume loss comparisons were made by grouping NAA10 and NAA15 probands together, rather than analyzing them separately, due to small sample sizes. Further experimentation with larger samples should be planned to better discriminate the effects of cortical thinning in NAA10 and NAA15 separately as opposed to individually.

The globus pallidus is both an anatomical and functional component of the basal ganglia that helps modulate movement and goal-oriented behavior [40]. Hyperintensities of the globus pallidus carry a broad differential diagnosis, including delayed or abnormal myelination, neoplasm, neurodegenerative disease, ischemia, metabolic disease, toxins, and infection [41]. Those with Ogden Syndrome were more likely to have hyperintensities of the globus pallidus than those with NAA15 related neurodevelopmental syndrome, potentially reflecting delayed myelination in this region. Additionally, those with hyperintensities of the globus pallidus were more likely to score lower on the motor portions of the Vineland. Decreased motor scores were also observed in probands with cerebellar hypoplasia, as opposed to those without [42]. Small sample sizes may be underlying the lack of statistical significance in both cases.

The olfactory bulb is a key component of the signal transduction pathway for olfaction, which continues to develop postnatally until the age of two [43]. There is preliminary evidence in rodent models that suggests olfaction also plays a role in child-parent socialization behaviors [44]. The increased incidence of olfactory bulb hypoplasia in NAA10-related neurodevelopmental syndrome versus NAA15-related neurodevelopmental syndrome could help explain poorer socialization in that cohort.

White matter is the network of fibers that connects grey matter in the brain, allowing for communication, feedback loops, and parallel processing to occur [45]. Those with white matter hypoplasia or abnormalities of their white matter scored significantly lower than those without white matter abnormalities on the written sub-domain (reading and writing skills) portion of the Vineland exam. Current literature suggests white matter volume is correlated with the acquisition of language development in various disease states [46, 47]. Written language requires the coordination of several cortical and sub-cortical areas, so proper flow of information is required for proper written language learning [48]. However, the cohort with diminished white matter volume was significantly smaller than the normal white matter cohort, and additional studies are needed to determine if white matter hypoplasia is associated with NAA10 and NAA15 related neurodevelopmental syndromes and if there is a dosimetric response correlating white matter volume with written language abilities.

Changes between scans were not observed in any probands with NAA15 variants in our cohort (n=7 NAA10, 3 NAA15) **(Supplementary Table 2)**. This is interesting to note, given that NAA10 mutations are generally regarded as causing more severe phenotypes. These findings may serve as a preliminary suggestion that NAA10 variants cause more progressive phenotypic changes on imaging, compared to a more stable phenotype seen in NAA15 variants.

One of the limitations of this study is the small sample size, which may have contributed to the lack of significance of many of our findings. Another limitation of the study is the difference in time between when behavioral testing and imaging was performed. Further experimentation should aim to align behavioral testing with the time of imaging to ensure that there is no change in between when the two take place. Additionally, only a single board certified neuroradiologist analyzed the images, so it is possible that there might be different interpretations between multiple neuroradiologists. Furthermore, due to the rare nature of the disease, participant scans were collected from institutions across the world. This increase in the diversity of participant backgrounds which helps broaden the applicability of results. However, it also increases the likelihood for minor differences in MRI protocol that may exist on an institution-by-institution basis. Additional prospective studies could possibly address each of these issues to cross-validate these results. Lastly, 3D volumetric analysis of the imaging data was not performed. This prevents comparisons of cortical and sub-cortical thickness that could have helped differentiate if decreases in brain volume were due to brain growth being delayed compared to the general population or if there are neurodegenerative processes occurring.

## CONCLUSIONS

Neuroanatomic deformities were generally more pronounced in probands with NAA10 variants, compared to those with variants in NAA15, consistent with previous data suggesting the former produces more severe phenotypes than the latter. More significant radiologic findings seem to correlate with poorer performance on Vineland testing, most notably with regards to motor skills. Some neuroanatomical deformities showed evolution over time, but additional imaging and testing will need to be performed to determine their significance.

## Data Availability

All data are deidentified to protect patient privacy, and the underlying data cannot be shared due to these same privacy restrictions.

## SUPPLEMENTAL INFORMATION (available upon request)

**Supplemental Table 1.** Proband Mutations and Syndromes by Number.

**Supplemental Table 2.** Full Patient Neuroimaging Data.

**Supplemental Table 3.** Neuroimaging Data for Probands with Changes Between Scans.

**Supplemental Table 4.** Clinical manifestations and Vineland-3 scores of 5 participants in our cohort with severe brain abnormalities as per their MRI results.

**Supplemental Note 1.** Patient 25 Case Summary and Occupational Therapy Evaluation Notes

**Supplemental Note 2.** Further Information on Probands with Changes Between Scans.

## Ethical Approval

Both oral and written patient consent were obtained for research and publication, with approval of protocol #7659 for the Jervis Clinic by the New York State Psychiatric Institute - Columbia University Department of Psychiatry Institutional Review Board.

